# Coding Complete Genome Sequences of Twenty-three SARS-CoV-2 Strains Isolated in the Philippines

**DOI:** 10.1101/2020.09.29.20203695

**Authors:** John Mark Velasco, Piyawan Chinnawirotpisan, Khajohn Joonlasak, Wudtichai Manasatienkij, Angkana Huang, Ma.. Theresa Valderama, Paula Corazon Diones, Susie Leonardia, Ma.Leanor Timbol, Fatima Claire Navarro, Vicente Villa, Henry Tabinas, Domingo Chua, Stefan Fernandez, Anthony Jones, Chonticha Klungthong

**Affiliations:** Department of Virology, U.S. Army Medical Directorate – Armed Forces Research Institute of Medical Sciences, Bangkok, Thailand; V Luna Medical Center, Armed Forces of the Philippines Health Service Command, Quezon City, Philippines; University of the Philippines Manila, Ermita, Manila, Philippines

## Abstract

Here, we report the coding complete genome sequences of 23 severe acute respiratory syndrome coronavirus 2(SARS-CoV-2) strains from the Philippines. Sequences were obtained from nasopharyngeal and oropharyngeal swabs from COVID-19 positive patients. Mutation analysis showed the presence of the D614G mutation in the spike protein in 22 of 23 genomes.

---

Severe acute respiratory syndrome coronavirus 2 (SARS-CoV-2) belonging to the family *Coronaviridae* and genus *Betacoronavirus* is the causative agent of COVID-19(1). In the Philippines, the first confirmed SARS-CoV-2 case was reported on 30 January 2020(2) and as of 31 Aug 2020, more than 200,000 cases had been reported(3). Here, we announce the coding complete genome sequences of 23 SARS-CoV-2 strains collected from 3 April-18 July 2020 from COVID-19 RT-PCR positive Filipino patients(4). Viral RNA was extracted from nasopharyngeal/oropharyngeal swabs using QIAamp viral RNA Mini kit (Qiagen) and used as template for amplicon sequencing using ARTIC SARS-CoV-2 V3 primers(4). DNA libraries were constructed and multiplexed using Illumina DNA prep, and then pooled before sequencing. Sequencing was performed on MiSeq using the MiSeq reagent kit V 2(Illumina). Reference mapping was performed by BWA-MEM aligner using Wuhan-Hu-1 genome sequence(GenBank Accession number NC_045512.2) as the reference sequence(5). Primer regions trimming and call variants (Q ≥25) were performed using iVAR v.1.2.2(6) and samtools (7), respectively. Consensus sequences were generated by iVAR v.1.2.2(6) (Q ≥25 and depth of coverage ≥10). Gaps, deletion, and ambiguous bases were identified and confirmed by genome-guided assembly with the reference sequence(NC_045512.2) using Trinity v2.8.5(8) and Sanger sequencing. Lineage and clade were determined using Pangolin v.2.0.4(9), GISAID clade nomenclature(10), and phylogenetic analysis(11, 12, 13). We used MEGA 7.0(14) to examine nucleotide and amino acid substitutions. All tools were run with default parameters. Raw reads yielded a total of 8.4 gigabases (94% of the clusters with Phred quality score (Q)≥30). The length of reads obtained were 35-251 nucleotides and the range of number of reads obtained per library was 0.87 – 1.48 million reads. Consensus sequences lengths were similar to the Wuhan-Hu-1 strain(NC_045512.2) with mean coverage ranging from 2,361X to 7,177X.

Table 1 shows strain details and mutations. 20/23 genomes collected between 25 June-18 July 2020 were classified under clade GR/lineage B.1.1, a major lineage frequently found in Europe and exported to the rest of the world(9, 10). Two genomes(2/23) from 6 and 7 July 2020 were classified under clade GR/lineage B.1.1.28, found in Brazil(89%), UK(8%), and China(2%) (https://cov-lineages.org/lineages/lineage_B.1.2.html). A single genome (1/23) was classified under clade O/lineage B.6, a global lineage mostly found in Singapore and India (https://cov-lineages.org). Four amino acid substitutions (ORF1b-P314L,S-D614G,N-R203K,and N-G204R), unique to clade GR (both lineages B.1.1 and B.1.1.28), were seen in all our sequences in this clade. Two sequences of lineage B.1.1 had a 21-nucleotide deletion in the N gene region. Five substitutions (ORF1a-R226K,ORF1a-E1126K,ORF1a-L3930F,S-V1176F,and ORF7b-V21I) were observed uniquely in 2 sequences of lineage B.1.1.28.

**Table 1.**
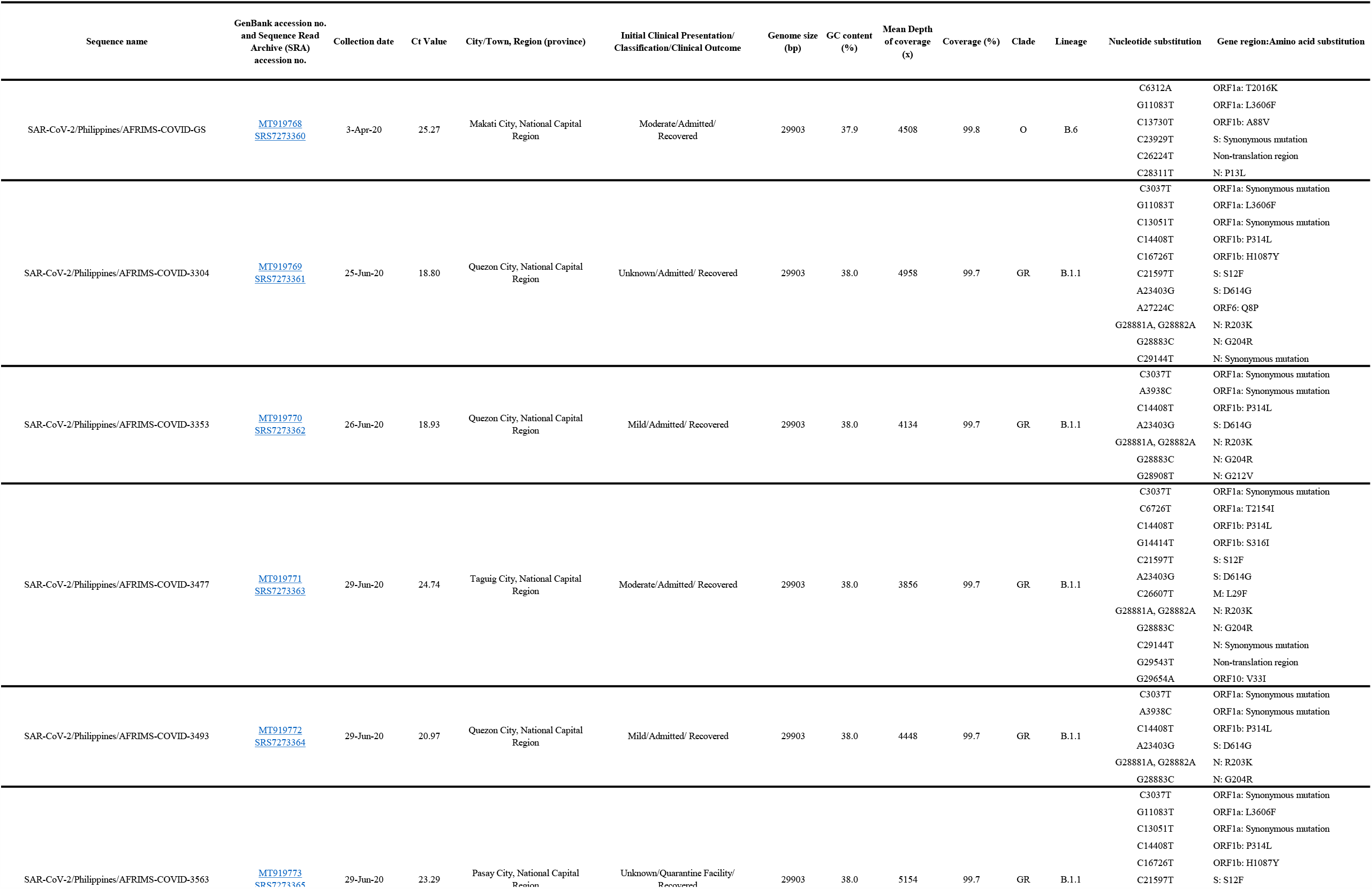
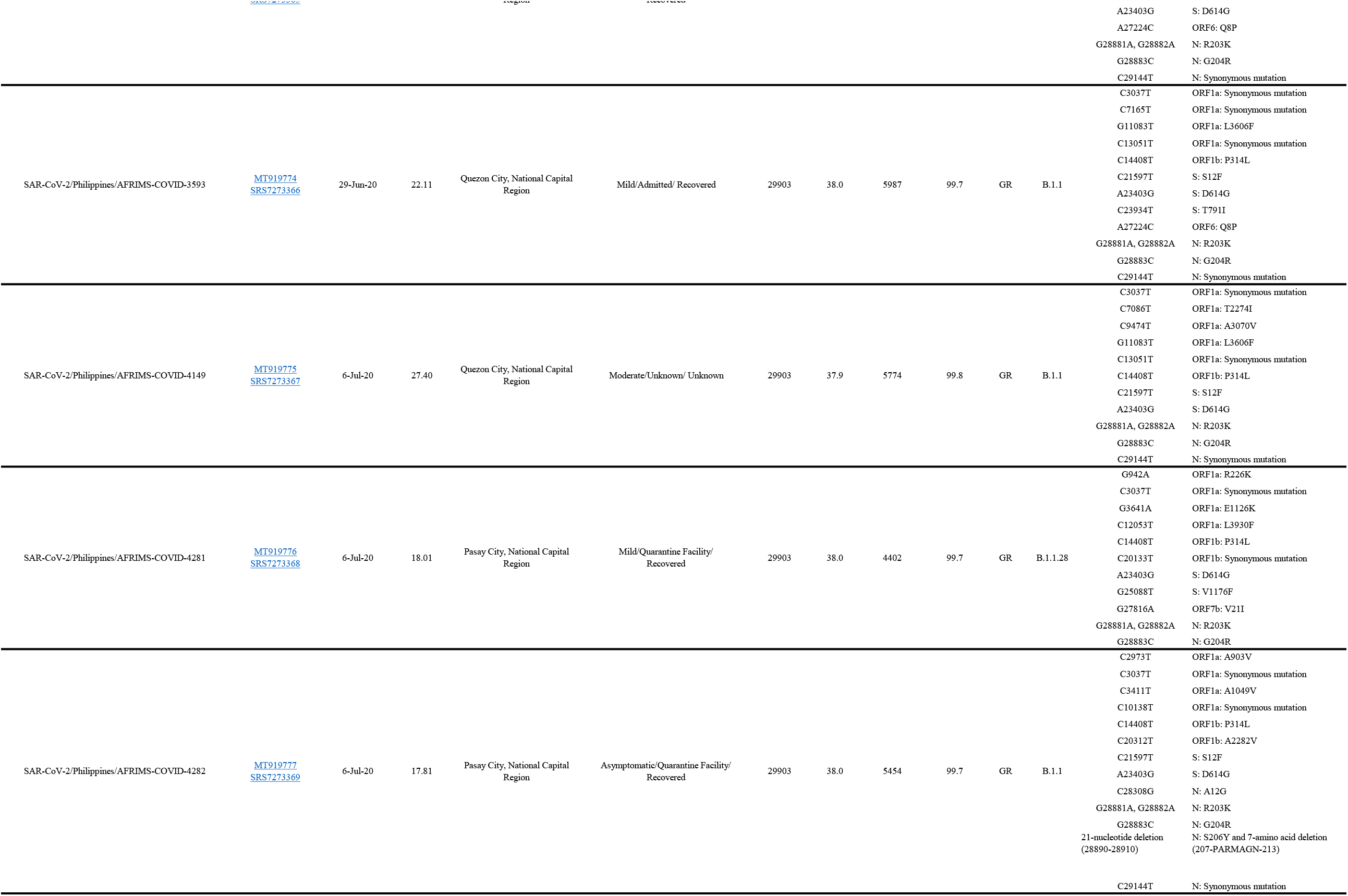
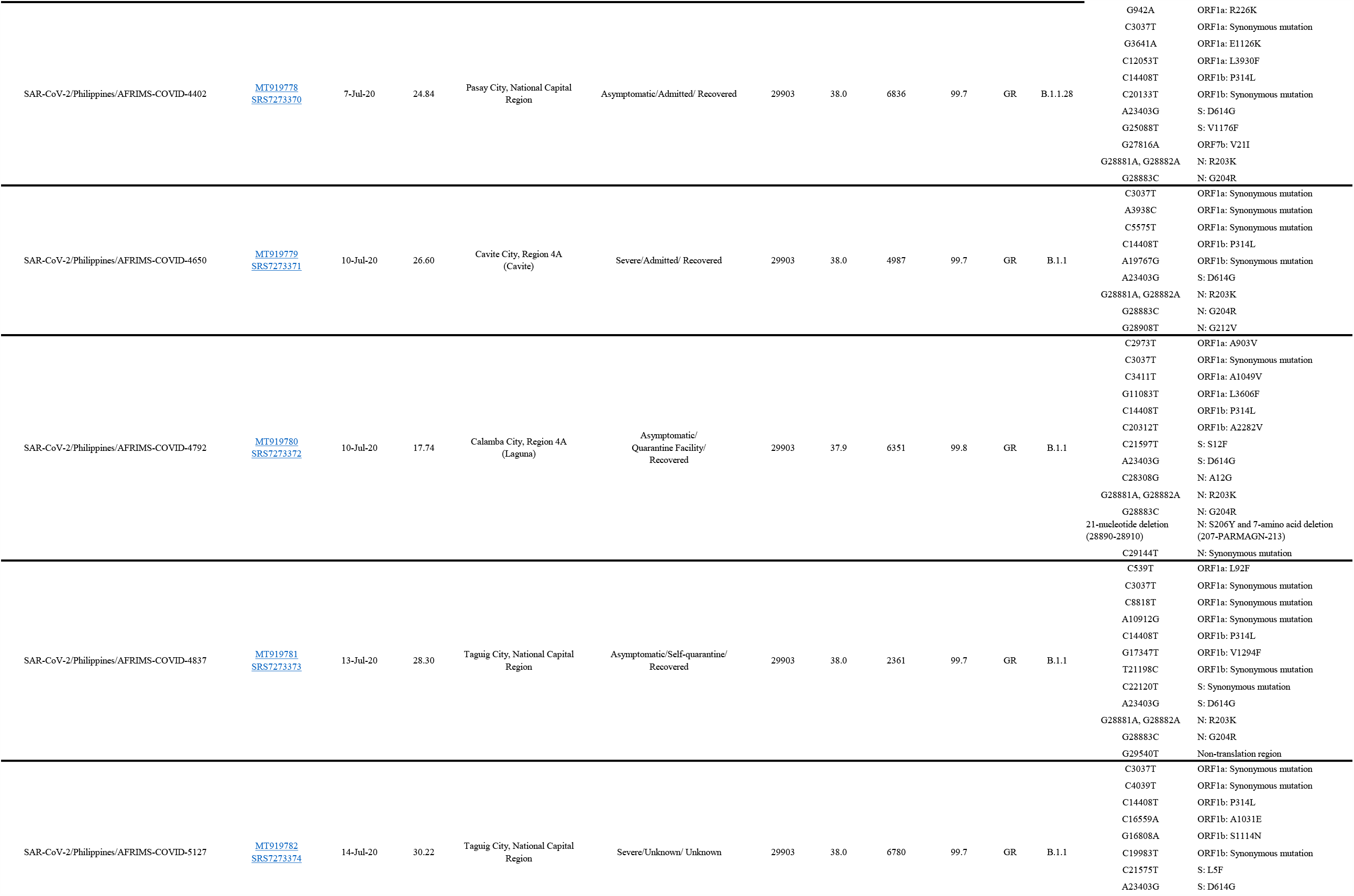
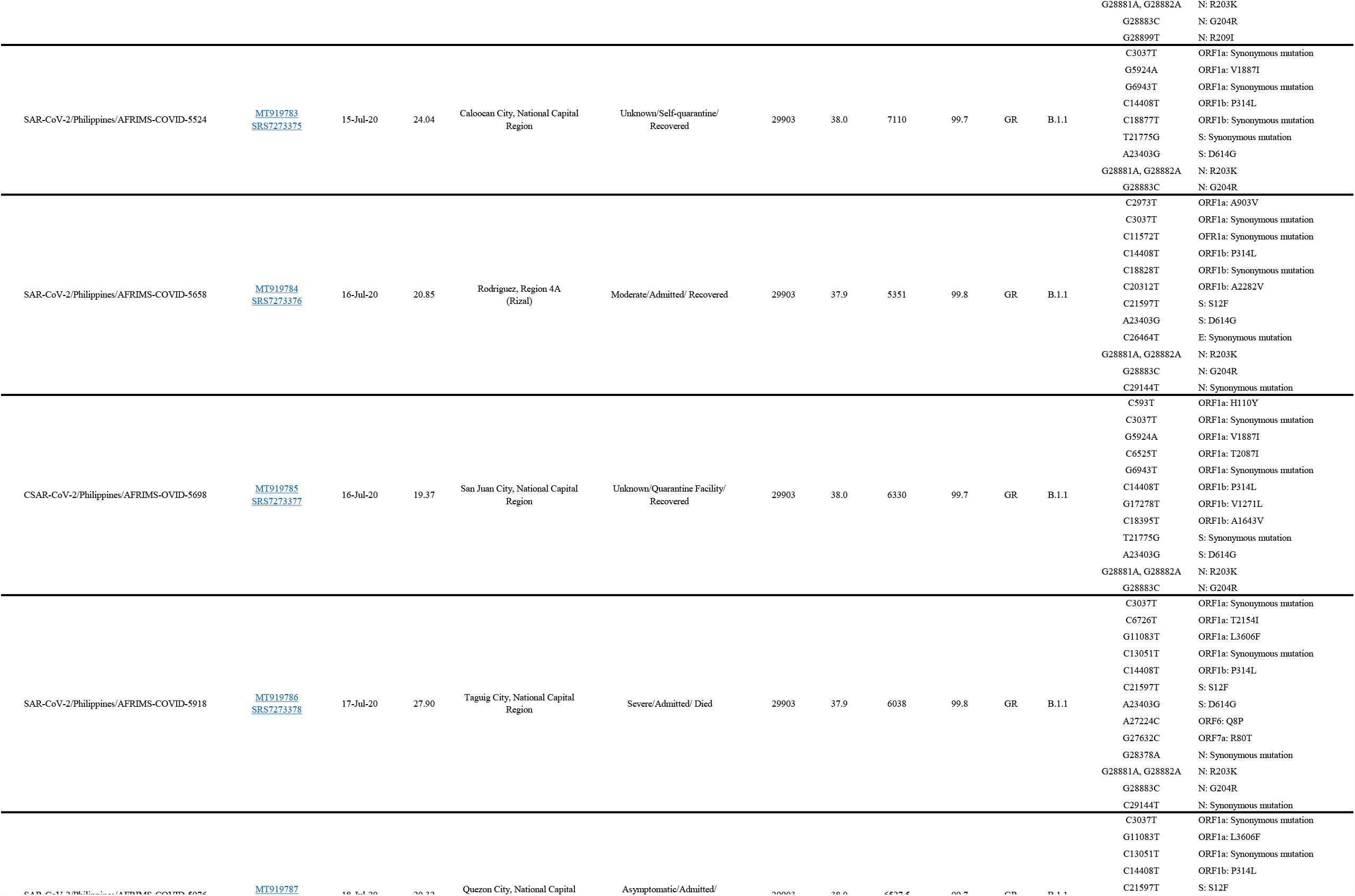
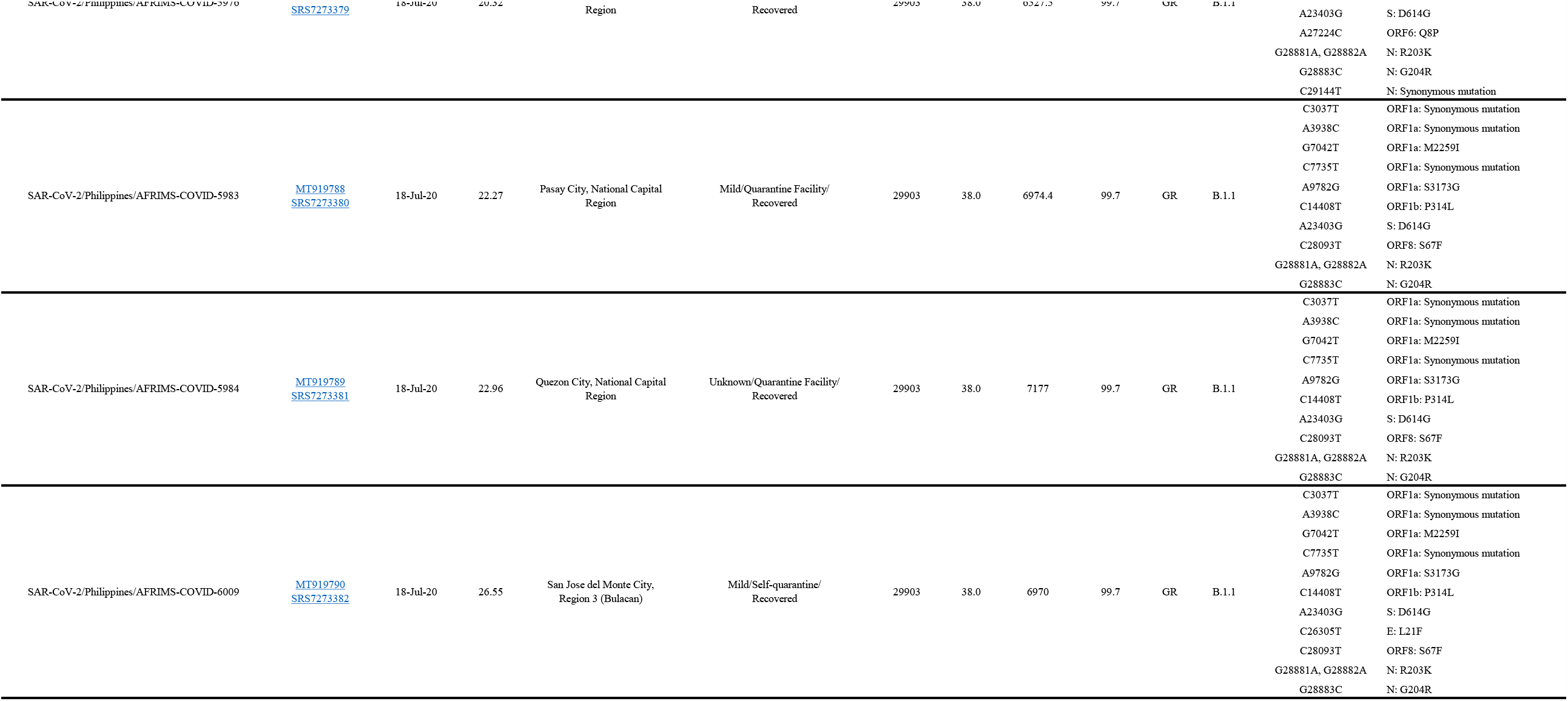
Genome features of twenty-three strains of SARS-CoV-2 from the Philippines.

None of the Philippine SARS-CoV-2 sequences in GISAID (accessed 22 August 2020) with collection date before 25 June 2020(15, 16) contained the D614G mutation. From the same set of data, lineage B.1.1 containing D614G was found in a sample collected on 25 June 2020. This observation coincides with our findings that the D614G mutation was observed from samples collected in June 2020(16). Our findings show the presence of lineages B.6, B.1.1 and B1.1.28, with the latter being first reported in the Philippines in this study.

The D614G mutation is currently the most prevalent variant worldwide and is associated with higher viral RNA levels and titers of pseudo viruses(17). In this study, all patients had no history of travel outside the country and acquired the disease in the National Capital Region, Region 3 and 4A(Table 1), providing evidence of local transmission. Multiple lineages and strains could have been introduced by travelers and Filipino repatriates(18). The D614G mutation is replacing and rapidly surpassing in prevalence the original strain circulating prior to June 2020 and may partially explain the rapid rise of cases in the Philippines.

## Data Availability

The SARS-CoV-2 genomes from the Philippines were deposited in the GenBank database (accession nos. MT919768-90). The raw reads have been deposited in the NCBI Sequence Read Archive (SRA accession nos. SRS7273360-82). The Bio Project accession no. is PRJNA659293. The Bio Sample accession nos. are SAMN15903138-60.

https://www.ncbi.nlm.nih.gov/nuccore/MT919768.1?report=fasta

https://www.ncbi.nlm.nih.gov/sra/?term=SRS7273360

https://www.ncbi.nlm.nih.gov/bioproject/?term=+PRJNA659293

https://www.ncbi.nlm.nih.gov/biosample/?term=SAMN15903138

## Acknowledgments

The authors acknowledge the support of the Department of Research and Training, Department of Pathology and Laboratories and Hospital Infection Control Committee of the V. Luna Medical Center (Quezon City, Philippines) for support with specimen collection. Informed consent for was obtained from the patients for SARS-CoV-2 testing. Sample collection was considered as a public health effort and did not require an Institutional Review Board approved human use protocol. This study was funded by the Armed Forces Health Surveillance Branch (AFHSB) and its Global Emerging Infections Surveillance (GEIS) Section, USA under grant number P0128_20_AF_13 for FY 2020. Material has been reviewed by the Walter Reed Army Institute of Research. There is no objection to its presentation and/or publication. The opinions or assertions contained herein are the private views of the author, and are not to be construed as official, or as reflecting true views of the Department of the Army or the Department of Defense. The investigators have adhered to the policies for protection of human subjects as prescribed in AR 70–25.

